# Predicting of COVID-19 Confirmed Cases in Different Countries with ARIMA Models in 2020

**DOI:** 10.1101/2020.03.13.20035345

**Authors:** Tania Dehesh, Heydar Ali Mardani-Fard, Paria Dehesh

## Abstract

The epidemic of a novel coronavirus illness (COVID-19) becomes as a global threat. The aim of this study is first to find the best prediction models for daily confirmed cases in countries with high number of confirmed cases in the world and second to predict confirmed cases with these models in order to have more readiness in healthcare systems. This study was conducted based on daily confirmed cases of COVID-19 that were collected from the official website of Johns Hopkins University from January 22^th^, 2020 to March 1^th^, 2020. Auto Regressive Integrated Moving Average (ARIMA) model was used to predict the trend of confirmed cases. Stata version 12 and R version 3.6.2 were used. Parameters used for ARIMA were (2,1,0) for Mainland China, ARIMA(1,0,0) for South Korea, and ARIMA(3,1,0) for Thailand. Mainland China and Thailand were successful in haltering COVID-19 epidemic. Investigating their protocol in this control like quarantine should be in the first line of other countries’ program

## Background

A novel Virus belongs to Corona virus ‘s family that has been transmitter from animal to human and was recognized in Wuhan, China, in December 2019. This can cause serious illness and death^1^. It has since been identified as a zoonotic coronavirus, similar to severe acute respiratory syndrome coronavirus (SARS-CoV) and Middle East Respiratory Syndrome Coronavirus (MERS-CoV), and was named 2019- nCoV^2^. A total number of 4515 cases including 106 deaths were confirmed on 27^th^ of January 2020^3^. A local seafood market in Wuhan was visited by many cases in the initial research, and it is indicated that a common-source zoonotic exposure may cause this new illness ^4^ The prevalence scope of this disease is unclear, because at present the prevalence of this disease is so dynamic^1^. There is obvious variation among countries in epidemiological surveillance and detection capacity for suspected cases. ^5^ Several cases of COVID-19 infections were also reported outside China, in other Asian countries, the United States, France, Australia, and Canada. In this situation when the illness does not have any specific treatment, the prevention of disease and preparation in healthcare services is very important .Modeling and future forecast of daily number of confirmed cases can help the treatment system in providing services for the new patients .The statistical prediction models could be helpful in forecasting and controlling this global epidemic threat. Here in this study, Auto Regressive Integrated Moving Average (ARIMA) model could be useful to predict the confirmed cases of COVID-2019. This model has more ability compared to some prediction models such as wavelet neural network (WNN) and the support vector machine (SVM) in prediction of natural disasters^6^. The global geographic regions in this study are according to World Health Organization (WHO) classification of regions. The data of countries with high number of confirmed cases analyzed in this study was based on WHO regions. For each country, the best ARIMA model is identified, and then 17 future days is predicted. The daily confirmed cases data of COVID-2019 from January 22^th^, 2020 to March 1^th^, 2020 were collected from the official website of Johns Hopkins University and were used to build these models. The aim of this study is first to find the best predicting models for confirmed cases in countries with high number of confirmed cases in the world and second to predict confirmed cases with these models. These models can help in predicting patients in near future to have better treatment staff preparedness in these countries.

## Methods

### ARIMA models

A total number of 41 (from January 22^th^, 2020 to March 1^th^, 2020) days were collected to develop ARIMA model.

The ARIMA models are important techniques in time series analysis that could be used in auto correlated data analysis. These models include autoregressive (AR) model, moving average (MA) model, and seasonal autoregressive integrated moving average (SARIMA) model ^7^.

The parameters of ARIMA model is as follows: (p, d, q)(P,D,Q)S generally, p refers to the order of auto- regression, d refers the degree of trend difference, q refers to the order of moving average, P refers to the seasonal auto-regression lag, D refers to the degree of seasonal difference, Q refers to the seasonal moving average, S refers to the length of the cyclical pattern^8^.

Before analyzing, time series must become station-based on mean and variance. The Augmented Dickey-Fuller (ADF) is used ^9^ in recognizing stationary in the mean and Box Cox test in recognizing whether the time series is stationary based on variance or not. Log transformation and differences are remedy approaches to stabilize the time series for variance and mean, respectively ^10^. Seasonal differences were used to stabilize the series from seasonality trend. The initial number of ARIMA model was guessed through autocorrelation function (ACF) graph and partial autocorrelation (PACF) graph.

All the models that passed the residual test (normality and stationary in variance) were compared using Akaike information criterion (AIC). The model which has the least AIC was selected as the best model. The methodology of current study was based on a previous study as reference ^11^. Microsoft Excel 2016 was used to build the database of daily COVID-19 in the world and STATA version 12 software was adopted to develop the ARIMA model. R version 3.6.2 software was used for scatter plot and smoothed curve. The statistical significance level was set at 0.05.

## Ethics

Since no primary data collection was undertaken, no patient or public was involved; no formal ethical assessment or informed consent was required. All data were collected from the official website and all data were fully anonymized.

## Results

Figure 1, 2 and 3 show scatter plots with smoothed curves of confirmed, death and recovered cases of COVID-19 in different countries. Those figures show overview of the disease trend in countries that affected by the disease.

Table 1 shows the ACF and PACF plots used for choosing the model parameters. The reporting of these ACF and PACF showed that confirmed cases of COVID-2019 were not influenced by the seasonality. According to these plots the P and Q parameters of ARIMA models were guessed. Then the guess models were compared according to AIC value.

**Table 1.**
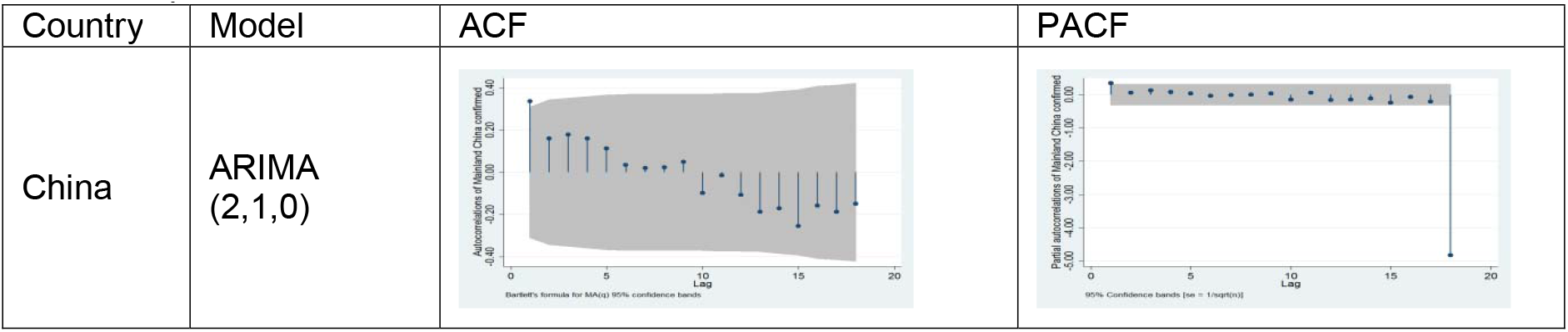
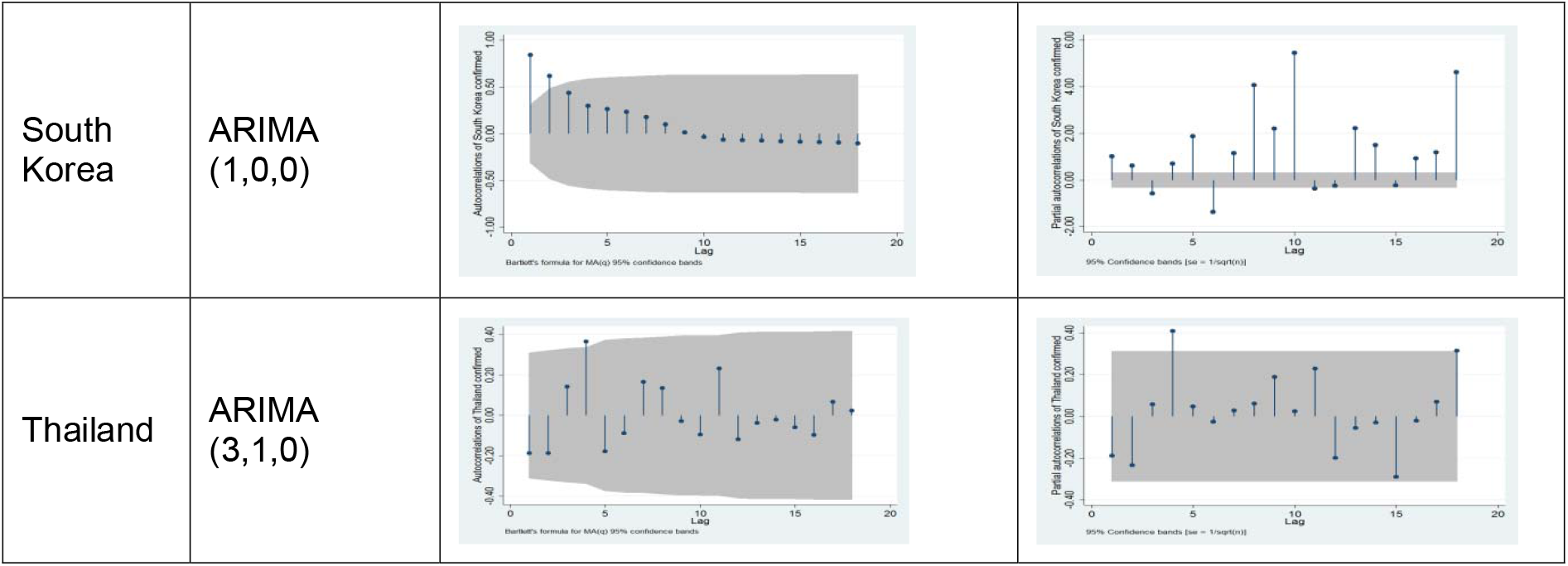
The best ARIMA models for forecasting number of daily confirmed cases according to ACF and PCF plots

The final models that were reported in table 2 had the lowest AIC values. Table 2 shows the forecast plots of ARIMA models for different countries with high number of confirmed cases according to WHO regions. The closes of predicted plots with actual confirmed data could be observed in these plots. This shows the precision of models in forecasting.

**Table 2.**
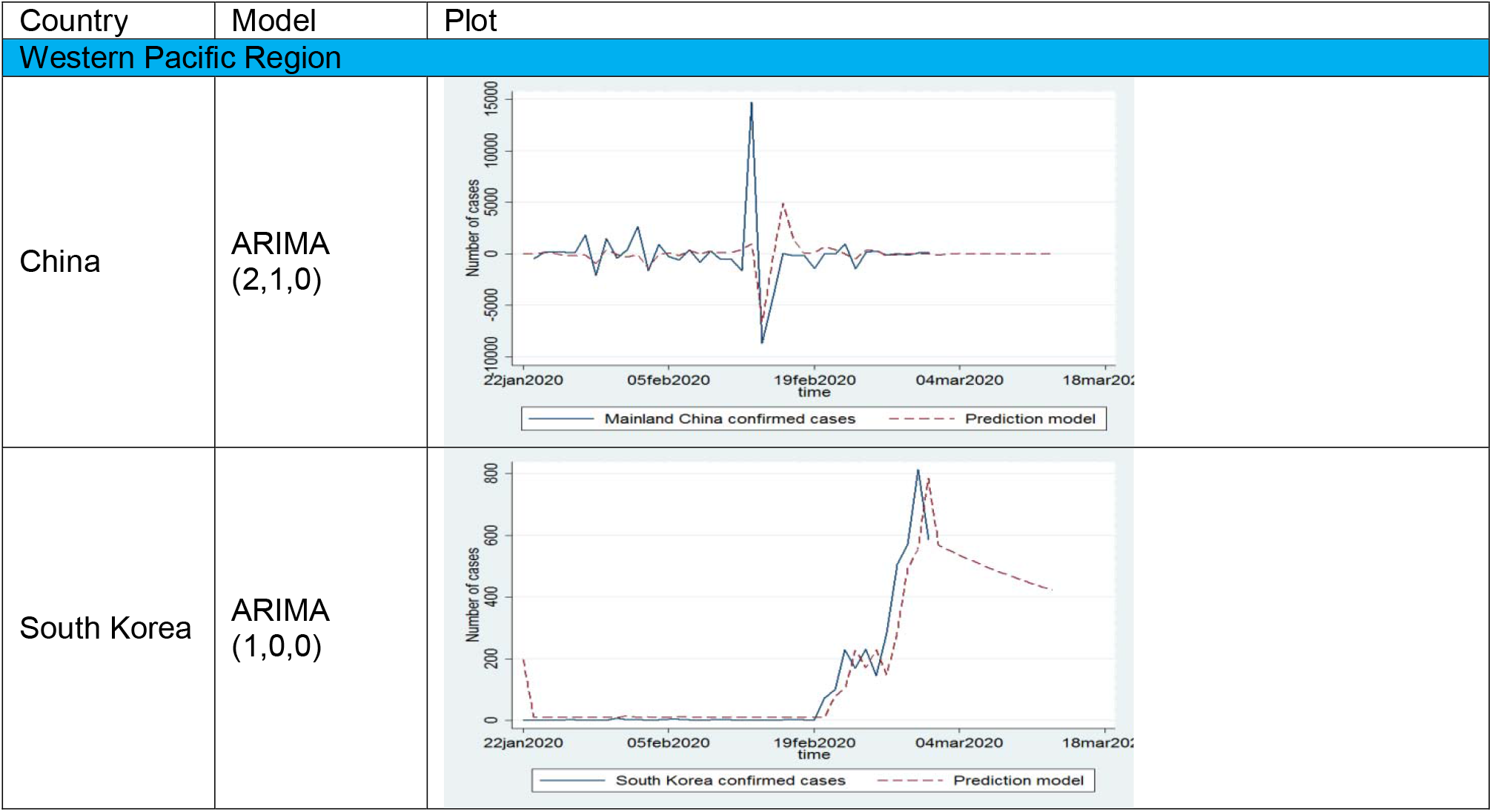
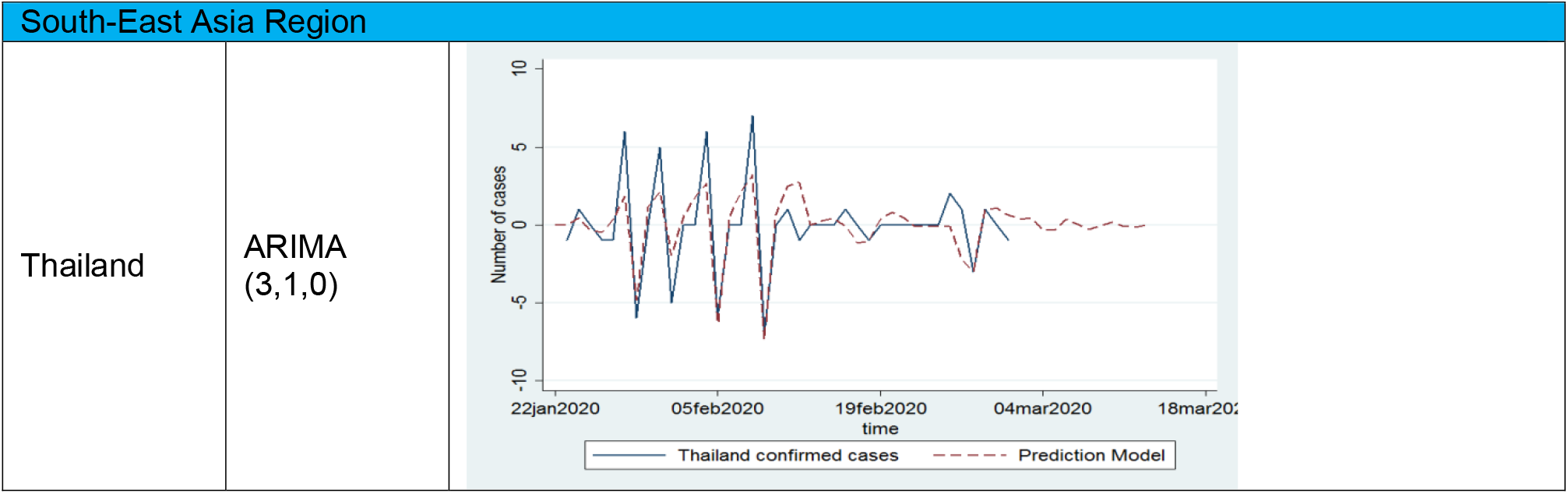
The plot of actual confirmed cases with the prediction result of ARIMA models in countries with high confirmed cases in different world regions (WHO regions)

Table 3 shows the forecast for 17 days (from 5^th^ of March until 21^th^ of March) with 95% confidence interval for different countries. This table shows that China may have stable trend after 6^th^ of March. South Korea presented a downward trend after 15^th^ of March. This hope is strong that this trend remains stable in near future in this country. Thailand almost controlled the epidemic and had zero or one confirmed cases daily. Iran and Italy did not have a stable trend in these 17 days.

## Discussion

The confirmed cases trend in China may become stable. South Korea will have a stationary trend in near future. Thailand almost controlled and the confirmed cases for this country were frequently between zero and one. Iran and Italy had unstable trend.

China ‘s confirmed cases showed stationary trend after 6^th^ of March. This result is in accordance with the result of previous study that predicted China ‘s epidemic to end up after March 10^th^, 2020^12^. This may be due to sever control and quarantine that were performed in China. This fact shows that quarantine worked well to reduce human exposure and control this epidemic. In another study on Chinese data the trend of confirmed cases in China during 10th–24th of January, 2020 was followed by exponential function ^13^.

This study confirmed that South Korea had an almost moderate trend that became decreasing after 15^th^ of March.

Iran did not have stationary trend even after 15^th^ of March. In a study according to the passengers that came from other countries such as UAE, Lebanon, and Canada the prevalence of cases in Iran estimated about 18,300 cases assuming an outbreak duration of 1.5 months in the country^14^. This paper was published online on 22.2.2020. Therefore Iran is at the beginning of the outbreak and the unstable trend was acceptable.

Thailand had the best control system in the exposure of this epidemic. Checking their program could be the best guide for future epidemics.

Although more data are needed to have a more detailed prevision, these models could be helpful in predicting future confirmed cases if the spread of the virus did not change very strangely. As we know this virus is novel and have the ability to be transmitted severely. This ability may affect all the predictions, but to our knowledge and for the time of writing, these models are the best.

## Conclusions

Mainland China and Thailand were successful in haltering COVID-19 epidemic. South Korea was also shown decreasing trend and may control this epidemic in near future. Investigating their protocol in this control like quarantine should be in the first line of other countries’ program

## Data Availability

Johns Hopkins University Center for Systems Science and Engineering, 2019. https://github.com/CSSEGISandData/COVIDQ3 19/blob/master/time_series/time_series_2019-ncov-Confirmed.csv

## Data references

Johns Hopkins University Center for Systems Science and Engineering, 2019. https://github.com/CSSEGISandData/COVIDQ319/blob/master/time_series/time_series_2019-ncov-Confirmed.csv

## References

1. Paules CI, Marston HD, Fauci AS. Coronavirus infections—more than just the common cold. JAMA. 2020;323(8):707–708.

2. Liu Y, Gayle AA, Wilder-Smith A, Rocklöv J. The reproductive number of COVID-19 is higher compared to SARS coronavirus. Journal of Travel Medicine. 2020.

3. Jung S-m, Akhmetzhanov AR, Hayashi K, et al. Real-Time Estimation of the Risk of Death from Novel Coronavirus (COVID-19) Infection: Inference Using Exported Cases. Journal of Clinical Medicine. 2020;9(2):523.

4. Huang C, Wang Y, Li X, et al. Clinical features of patients infected with 2019 novel coronavirus in Wuhan, China. The Lancet. 2020;395(10223):497–506.

5. Niehus R, De Salazar PM, Taylor A, Lipsitch M. Quantifying bias of COVID-19 prevalence and severity estimates in Wuhan, China that depend on reported cases in international travelers. medRxiv. 2020.

6. Zhang Y, Yang H, Cui H, Chen Q. Comparison of the Ability of ARIMA, WNN and SVM Models for Drought Forecasting in the Sanjiang Plain, China. Natural Resources Research. 2019:1–18.

7. Fattah J, Ezzine L, Aman Z, El Moussami H, Lachhab A. Forecasting of demand using ARIMA model. International Journal of Engineering Business Management. 2018;10:1847979018808673.

8. Wei W, Jiang J, Liang H, et al. Application of a combined model with autoregressive integrated moving average (ARIMA) and generalized regression neural network (GRNN) in forecasting hepatitis incidence in Heng County, China. PloS one. 2016;11(6).

9. Cao S, Wang F, Tam W, et al. A hybrid seasonal prediction model for tuberculosis incidence in China. BMC medical informatics and decision making. 2013;13(1):56.

10. Cheung Y-W, Lai KS. Lag order and critical values of the augmented Dickey–Fuller test. Journal of Business & Economic Statistics. 1995;13(3):277–280.

11. Wang Y-w, Shen Z-z, Jiang Y. Comparison of ARIMA and GM (1, 1) models for prediction of hepatitis B in China. PloS one. 2018;13(9).

12. Li Q, Feng W. Trend and forecasting of the COVID-19 outbreak in China. arXiv preprint arXiv:200205866. 2020.

13. Lai C-C, Shih T-P, Ko W-C, Tang H-J, Hsueh P-R. Severe acute respiratory syndrome coronavirus 2 (SARS-CoV-2) and corona virus disease-2019 (COVID-19): the epidemic and the challenges. International journal of antimicrobial agents. 2020:105924.

14. Tuite AR, Bogoch I, Sherbo R, Watts A, Fisman DN, Khan K. Estimation of COVID-2019 burden and potential for international dissemination of infection from Iran. medRxiv. 2020.

